# Problematic Internet Use in Children and Adolescents: Associations with psychiatric disorders and impairment

**DOI:** 10.1101/19005967

**Authors:** Anita Restrepo, Tohar Scheininger, Jon Clucas, Lindsay Alexander, Giovanni A. Salum, Kathy Georgiades, Diana Paksarian, Kathleen R. Merikangas, Michael P. Milham

**Affiliations:** Healthy Brain Network, Child Mind Institute, New York, New York; MATTER Lab, Child Mind Institute, New York, New York; Department of Psychiatry, Universidade Federal do Rio Grande do Sul, Porto Alegre, Brazil; Department of Psychiatry and Behavioural Neurosciences, McMaster University, Canada; Genetic Epidemiology Research Branch, Intramural Research Program, National Institute of Mental Health, Bethesda, MD 20895, USA; Center for the Developing Brain, Child Mind Institute, New York, New York; Center for Biomedical Imaging and Neuromodulation, Nathan S. Kline Institute for Psychiatric Research, Orangeburg, New York

**Keywords:** Internet addiction, Pediatric, Depression, ADHD, Impairment

## Abstract

**Objective:** Here, we leveraged the ongoing, large-scale Child Mind Institute Healthy Brain Network, a transdiagnostic self-referred, community sample of children and adolescents (ages 5-21), to examine the associations between Problematic Internet Use (PIU) and psychopathology, general impairment, physical health and sleep disturbances.

**Methods:** A total sample of 564 (190 female) participants between the ages of 7-15 (mean = 10.80, SD = 2.16), along with their parents/guardians, completed diagnostic interviews with clinicians, answered a myriad of self-report questionnaires, and underwent physical testing as part of the Healthy Brain Network protocol.

**Results:** PIU was positively associated with depressive disorders (aOR = 2.34; CI: 1.18-4.56; p = .01), the combined subtype of ADHD (aOR = 1.79; CI: 1.08-2.98; p = .02), greater levels of impairment (Standardized Beta = 4.79; CI: 3.21-6.37; p < .01) and increased sleep disturbances (Standardized Beta = 3.01; CI: 0.58-5.45; p = .02), even when accounting for demographic covariates and psychiatric comorbidity.

**Conclusion:** The association between PIU and psychopathology, as well as its impact on impairment and sleep disturbances, highlight the urgent need to gain an understanding of mechanisms in order to inform public health recommendations on internet use in U.S. youth.

## 1. INTRODUCTION

The terms “problematic internet use”, “internet addiction”, “compulsive internet use” and “pathological internet use” have been used to refer to patterns of problematic behavior associated with internet use. Regardless of the specific terminology employed, concerns about the potential harms of internet use are growing rapidly, particularly among mental health providers.^1–6^ There is a consensus that problematic internet use (PIU) is characterized by overuse of the internet with associated impairment(s) across various domains of functioning.^1–4,7,8^ Some have gone further, suggesting that PIU is a behavioral version of a substance-use disorder,^9–11^ while others have suggested that it is either an impulse-control disorder,^3,12,13^ or a subtype of obsessive-compulsive disorder,^14,15^ although empirical evidence for these designations is lacking.^8,10,16–20^ Regardless of whether PIU truly constitutes a form of addiction, there is a need to obtain a more complete empirical understanding of the contexts in which it arises in childhood and adolescence, as well as its unique contributions to socio-emotional and behavioral functioning.

Prevalence estimates of PIU typically range between 1-10%, although some estimates such as those in Asia have exceeded 25%.^3,21–30^Accumulating evidence has documented associations between PIU and the presence of one or more mental disorders.^4,31,32^ Depression has been consistently linked to PIU across studies measuring general internet use, smartphone use, and online gaming.^6,7,26,31–54^ Similarly, anxiety disorders, particularly social anxiety^26,37,43,44,47,55,56^ and Generalized Anxiety Disorder (GAD),^37,50,54^ have also been associated with PIU.^6,31,38–40,42,45,46,48,49,51,53^ PIU has been further shown to correlate with different components of anxious thought (i.e. fear of missing out, fear of negative evaluation) in the context of social anxiety.^57^ Though generally less consistent, associations between PIU and ADHD,^7,26,27,37,43,45,50,58,59^ personality disorders,^12,37^ obsessive-compulsive disorder,^7,31,37,46^ and either schizophrenia, psychotic symptoms or dissociative symptoms,^7,37,46,48^ have also been documented.

PIU has also been associated with impairment in various life domains, including problems with social relationships^4,7,30,43,60–62^ and professional endeavors.^7,12^ PIU has also been linked to lower reported life satisfaction and well-being.^40,43,63,64^ Consistent with the range of findings reported, individuals who sought online mental health help had higher scores on a broad impairment scale.^65^ PIU has also been associated with impairment in physiological and physical domains. For example, decreased physical fitness has been associated with PIU^49,66–70^ and physical activity was found to be a protective factor in the relationship between PIU and psychopathology.^49,71^ Additionally, there are links between PIU and sleep disturbances,^36,41,50,72^ insomnia,^42,73^ and lower sleep quality.^39,53,73^ The structure of the relationship between PIU, poor sleep, physical health and psychopathology is unclear, as some studies have suggested mediating or moderating effects amongst the different variables.^36,39,53^ It is unclear whether the impairment associated with PIU - both physiological and non-physiological - is independent from that caused by potentially comorbid psychological disorders present in individuals with PIU.^33,36,53,68,69^

While many prior studies have separately found associations between PIU and impairment and PIU and psychiatric disorders, the present work explores both multidiagnostic assessments and general impairment in a single, diverse sample of youth. Additionally, measures include both parent and self-reports, allowing for comparison between informants. Here, we leveraged the ongoing, large-scale Child Mind Institute Healthy Brain Network, a transdiagnostic self-referred community sample of children and adolescents (ages 5-21), to examine the associations between PIU and psychopathology. The aims were: 1) to examine associations between PIU and clinician diagnoses of a range of mental disorders in youth controlling for the effects of comorbidity; and 2) to assess whether PIU was associated with general impairment and a reduction in health behaviors including poor physical health and sleep disturbances.

## 2. MATERIALS AND METHODS

### 2.1 Sample/Design

Participants were recruited from the ongoing Healthy Brain Network (HBN) initiative that seeks to create and share a 10,000-participant biobank of data from children and adolescents ages 5-21 from the New York City area74. Data collected includes psychiatric, cognitive, behavioral, genetic, and lifestyle information as well as MRI and EEG neuroimaging. The HBN has two main collection sites -- one located on Staten Island and one in Midtown Manhattan. The study was approved by the Chesapeake Institutional Review Board. Written informed consent was obtained from adult participants. For participants younger than 18, written consent was obtained from their guardians and written assent obtained from the participant. Participants were excluded from participation if there were outstanding safety concerns, insufficient verbal abilities, an IQ lower than 66, and neurological concerns that could interfere with MRI and EEG interpretation. Participants were asked to suspend stimulant medication meant for treatment of ADHD on the days of testing to minimize potential confounds, though participants who did not suspend medication use were still included in the study and medication use was noted on the day of assessment.

As part of the HBN survey battery, participants and their parents/guardians completed a variety of age-based questionnaires assessing basic demographic characteristics, dimensional assessments of domains associated with mental health, substance use, socioeconomic status and internet use. Information about the participant’s race was collected from parent verbal report during a structured clinical history interview. For participants under the age of 11, a trained research assistant read and explained individual items and collected responses from participants.

### 2.2. Measures

#### 2.2.1 Schedule for Affective Disorders and Schizophrenia - Children’s Version (K-SADS)

Participants and their parents/guardians were independently administered the Schedule for Affective Disorders and Schizophrenia - Children’s Version (K-SADS)^75^ by a trained member of the clinical team. For participants under the age of eleven, clinicians determined whether the K-SADS would be administered to both child and parent or just parent based on verbal function and expected ability to tolerate the interview. Lifetime consensus diagnoses were based on DSM-5 diagnostic criteria using information from both the parent and child along with historical records and other HBN assessments^74^ when clarifications were needed.

#### 2.2.2 Internet Addiction Test (IAT)

Young’s IAT is a 20-item questionnaire ranked on a five-point Likert-type scale.^76^ Both parent and self-reported versions were used to assess PIU. The total score resulted from summing all 20 items on the questionnaire, resulting in possible scores ranging from 0-100. The internal consistency was adequate in the current sample with a cronbach’s alpha value of 0.91 for self-report and 0.95 for parent-report.

Based on previous work,^9,45,77^ a total sum score of 40 or above was categorized as Problematic Internet Use (PIU). To check the validity of this threshold, the value of 40 was increased. For subsequent analyses, participants were then categorized as either non-problematic (absence of PIU) or problematic internet users (presence of PIU) using the cut-off value of 40.

#### 2.2.3 The Barratt Simplified Measure of Social Status (BSMSS)

The BSMSS is a parent-reported measure of social status, which is used as a proxy for socioeconomic status (SES).^78^ The total sum score used in the current report is calculated by adding the scores for the total occupation and total education subscales, which are answered in regards to both guardians. For participants with a single caregiver, total scores were calculated based solely on that caregiver’s responses. Total scores were then divided into tertiles to determine low, middle and high SES, respectively, for the demographics analysis and included as a continuous covariate for the rest. Since the presence or absence of a secondary caregiver explained a large amount of the partial missing data in the BSMSS, an additional binary variable representing this factor was also included as a covariate in all analyses.

#### 2.2.4 Columbia Impairment Scale (CIS)

The CIS is a 13-item scale that assesses global functioning in domains of interpersonal relations, psychopathology, school performance, and use of leisure time.^79^ The total sum score of all items was used for both self and parent-report versions. Internal consistency was adequate in the current sample (cronbach’s alpha for parent-report was 0.85 and 0.81 for self-report).

#### 2.2.5 Physical Activity Questionnaire (PAQ)

The PAQ is a self-report questionnaire asking participants to recall their physical activity levels in various domains for the past seven days.^80^ The PAQ-C is administered to children ages 8-14 and the PAQ-A, modified by removing items referring to school recess, is administered to ages 14-19. The total score is created using the average of all items after the first question, which are all rated on a scale from one to five. Cronbach’s alpha was 0.88 for the PAQ-C and 0.87 for the PAQ-A.

#### 2.2.6 Sleep Disturbances Scale (SDSC)

The SDSC is a parent-reported 27-item questionnaire rated on a 5-point Likert type scale that assesses sleep disturbances in various domains (initiating and maintaining sleep, breathing, disorders of arousal, sleep-wake transition, excessive somnolence, and sleep hyperhidrosis).^81^ The total sum score of all items was used in the current report. Cronbach’s alpha in the current sample was 0.82.

#### 2.2.7 Body Composition Measures

Participant height and weight were also collected by a trained research assistant in order to calculate Body Mass Index (BMI). Electric impedance and reactance measures were collected using electrodes placed at four positions on the body: two on the right hand and two on the right foot. RJL Systems Bio-Impedance Analysis was used to calculate Fat Mass Index (FMI).

### 2.3 Analyses

Main analyses were completed using self-report measures (corresponding analyses using parent-report measures can be found in the supplementary material). Participants who did not complete one or more of the questionnaires required for the analyses were removed from the sample. Due to the small number of individual missing items in the remaining sample, complete cases analysis was used. All analyses were also run using the maximum number of complete datasets for each (thus resulting in differential sample sizes for each analysis) in order to determine whether results differed when analyses were run using the final sample (see Tables S7-S12). The final sample consisted of 564 (190 female) participants between the ages of 7-15 (mean = 10.80, SD = 2.16) out of the original sample of 2094 children aged 5-21. Regarding those excluded (n = 1,530), 1,310 were missing data due to key questionnaires included in the present work being introduced later in the protocol’s timeline, and the remaining 220 were missing a combination of demographic information, physical activity information, and/or KSADS administration. All statistical analyses were conducted using The R Project for Statistical Computing for Windows^82^ (version 3.5.2).

To examine demographic correlates of PIU, unadjusted prevalence odds ratios were calculated using the R package epiR^83^. Next, logistic regression was used to estimate three odds ratios (ORs) models: unadjusted ORs for associations between PIU and mental disorders, ORs adjusted for age, sex, SES, site, and single caregiver, as covariates, and adjusted ORs that also accounted for comorbid mental disorders. Third, linear regression was used to assess the relationship between PIU and impairment. We again estimated three sets of models: unadjusted, adjusted for age, sex, SES, site, and single caregiver, and further adjusted for mental disorders.. Finally, we estimated associations of PIU with physical health and sleep disturbances using linear regression. Again, we first estimated unadjusted associations, followed by adjustment for demographics, and finally adding adjustment for mental disorders.

## 3. RESULTS

### 3.1 Demographics

Although there were no significant differences in PIU by any of the demographic characteristics, PIU was more common in males than females, in those ages 10-12, and those in the high SES range. See Figure 1 and Figure S1 for the distribution of IAT scores across age groups and sex for self and parent-report, respectively.

**Figure 1:**
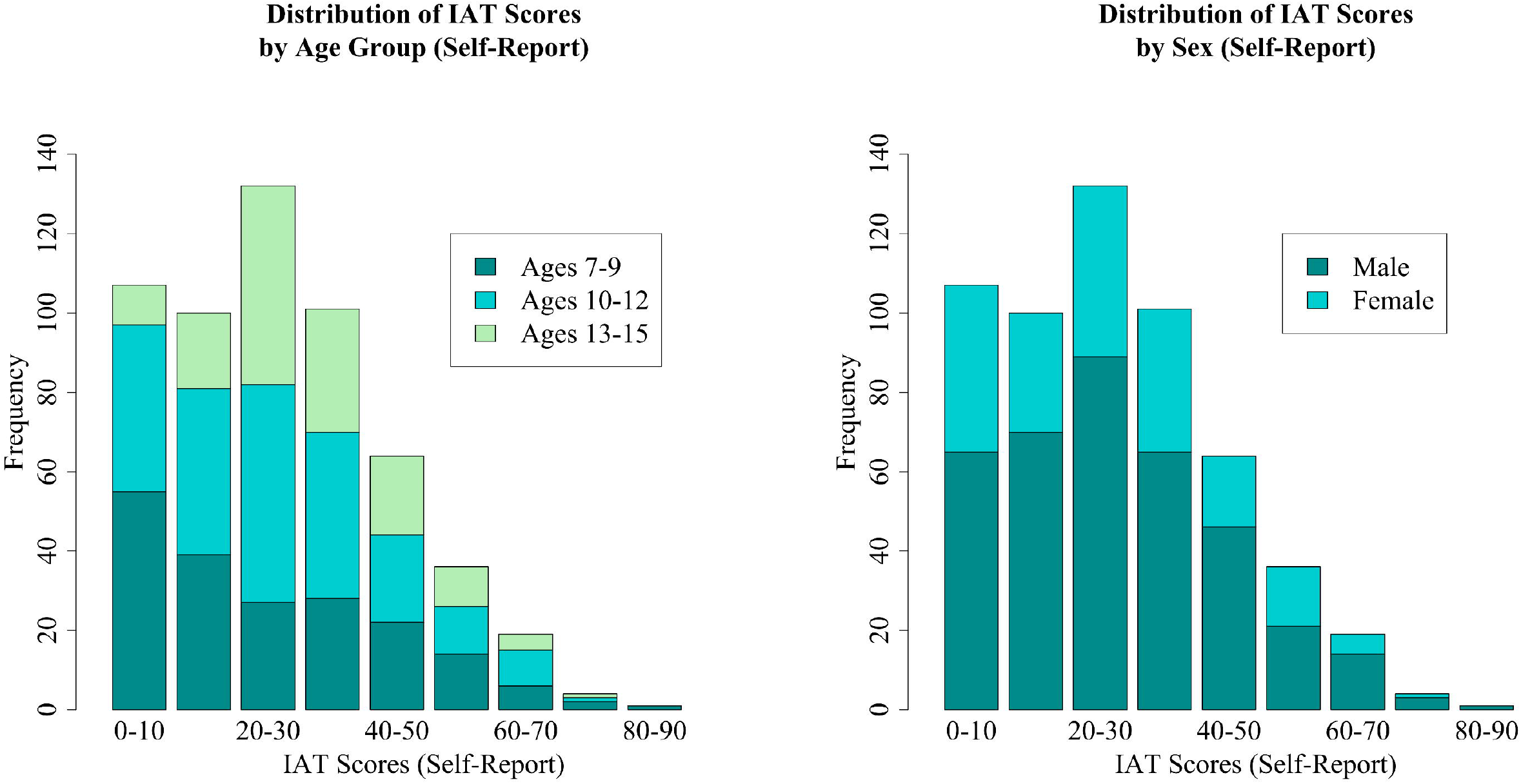
Self-reported Internet Addiction Test score distributions broken down by participant age group and sex.

### 3.2 Mental Disorders

Table 2 shows the rates and odds ratios for associations between PIU and psychiatric disorders. After adjustment for age, sex, SES, site, single caregiver and all other diagnoses of interest, there were significant positive associations between PIU and depressive disorders (aOR = 2.42; CI: 1.22-4.73; p = .01) and the combined subtype of ADHD (aOR = 1.89; CI: 1.13-3.17; p = .02).

**TABLE 1.** Number of participants within each demographic category who scored below and above 40 on the IAT; unadjusted odds ratios for each category.

|  | PIU - Absent<sup>a</sup> | PIU - Present<sup>b</sup> | OR (95% CI) | *p* |
| --- | --- | --- | --- | --- |
| **Sex** |  |  |  |  |
| Male | 289 [51.24%] | 85 [15.07%] | 1.00 (ref) | - |
| Female | 151 [26.77%] | 39 [6.91%] | 0.88 (0.57-1.34) | 0.55 |
| **Age** |  |  |  |  |
| 7-9 | 149 [26.42%] | 45 [7.98%] | 1.00 (ref) | - |
| 10-12 | 181 [32.09%] | 44 [7.8%] | 0.80 (0.50-1.28) | 0.36 |
| 13-15 | 110 [19.50%] | 35 [6.21%] | 1.05 (0.64-1.74) | 0.84 |
| **SES** |  |  |  |  |
| Low SES | 52 [9.22%] | 14 [2.48%] | 1.00 (ref) | - |
| Middle SES | 85 [15.07%] | 38 [6.74%] | 1.66 (0.83-3.33) | 0.16 |
| High SES | 303 [53.72%] | 72 [12.77%] | 0.88 (0.47-1.67) | 0.70 |
| **Race** |  |  |  |  |
| Caucasian | 240 [42.55%] | 58 [10.28%] | 1.00 (ref) | - |
| African American | 52 [9.22%] | 21 [3.72%] | 1.67 (0.94-2.98) | 0.08 |
| Hispanic | 43 [7.62%] | 16 [2.84%] | 1.54 (0.82-2.91) | 0.19 |
| Asian | 11 [1.95%] | 3 [0.53%] | 1.13 (0.33-3.90) | 0.86 |
| Other Race | 94 [16.67%] | 26 [4.61%] | 1.14 (0.68-1.92) | 0.61 |
| **Site** |  |  |  |  |
| Staten Island | 243 [43.09%] | 72 [12.77%] | 1.00 (ref) | - |
| Midtown | 197 [34.93%] | 52 [9.22%] | 0.89 (0.60-1.33) | 0.57 |
| *Note: <sup>a</sup>(n=440 [78.01%]); <sup>b</sup>(n=124 [21.99%]).* |  |  |  |  |

**TABLE 2.** Prevalence rates and adjusted odds ratios (95% CI) for associations between DSM-5 disorders (lifetime prevalence) and Problematic Internet Use (PIU) determined by a score above or below a cut-off score of 40 on the Internet Addiction Test

| Diagnosis | PIU - Absent<sup>a</sup> (n [%]) | PIU - Present<sup>b</sup> (n [%]) | OR (95% CI) | *p* | aOR<sup>c</sup> (95% CI) | *p* | aOR<sup>d</sup> (95% CI) | *p* |
| --- | --- | --- | --- | --- | --- | --- | --- | --- |
| ASD | 61 [10.82%] | 25 [4.43%] | 1.57 (0.94-2.62) | 0.08 | 1.57 (0.92-2.63) | 0.09 | 1.47 (0.85-2.49) | 0.16 |
| Anxiety | 140 [24.82%] | 43 [7.62%] | 1.14 (0.75-1.73) | 0.55 | 1.16 (0.75-1.78) | 0.49 | 1.00 (0.59-1.66) | >0.99 |
| Depression | 31 [5.50%] | 19 [3.37%] | 2.39 (1.30-4.37) | <.001 | 2.41 (1.25-4.56) | 0.01 | 2.42 (1.22-4.73) | 0.01 |
| ADHD-C | 107 [18.97%] | 46 [8.16%] | 1.84 (1.20-2.80) | <.001 | 1.96 (1.25-3.06) | <.001 | 1.89 (1.13-3.17) | 0.02 |
| ADHD-I | 140 [24.82%] | 36 [6.38%] | 0.88 (0.57-1.35) | 0.55 | 0.81 (0.52-1.26) | 0.36 | 1.05 (0.63-1.75) | 0.86 |
| Social Anxiety | 49 [8.69%] | 14 [2.48%] | 1.02 (0.55-1.90) | 0.96 | 1.01 (0.52-1.88) | 0.97 | 0.94 (0.43-2.01) | 0.88 |

**Note:** <sup>a</sup>(n=440 [78.01%]); <sup>b</sup>(n=124 [21.99%]); <sup>c</sup>Adjusted for sex, age, SES, collection site, and single caregiver; <sup>d</sup>Adjusted for sex, age, SES, collection site, single caregiver and all diagnoses.

The IAT threshold score used to differentiate between problematic and non-problematic use was varied in order to test the validity of the score suggested by previous literature. When the threshold score was increased to 50, unadjusted odds ratios for self-reported PIU remained significant for ADHD combined type, though not for depressive disorders, and there was a statistically significant association between PIU and the ADHD inattentive type. These associations were no longer significant after the threshold score was increased to 60 (out of 100).

Of note, examination of rates of depression and the combined subtype of ADHD in participants that were excluded from the main analyses yielded marginally significant to significantly elevated odds ratios for depression (self report PIU (n = 461): 1.60 [0.92-2.79], p = .09; parent report PIU (n = 876): 4.22 [2.55-6.98], p < 0.001) and ADHD, combined presentation (parent report PIU (n = 876): 1.41 [0.98-2.01], p = .06; self report PIU: n.s.). These findings further support our primary findings and alleviate possible concerns about the impact of exclusion criteria.

### 3.3 Indices of Impact of PIU

Table 3 presents the associations between PIU and impairment assessed by self-reported CIS scores, physical fitness assessed by self-reported PAQ scores, BMI, FMI, and sleep disturbances assessed by parent-reported SDSC scores. After adjustment for demographic factors and psychiatric disorders, PIU was associated with greater levels of impairment (Standardized Beta = 4.79; CI: 3.21-6.37; p < .01). Although BMI was positively associated with PIU in the unadjusted model, no significant differences emerged after adjustment for demographic and psychiatric disorders (Standardized Beta = 0.67; CI: −0.23-1.58; p = .15). There was no significant association between PIU and our indicator of physical fitness or FMI (see Table 3). PIU was significantly associated with sleep disturbances in the adjusted models (Standardized Beta = 3.01; CI: 0.58-5.45; p = .02).

**TABLE 3.** Adjusted associations between Problematic Internet Use (PIU) with impairment, physical health, and sleep.

| Outcome | Model 1<sup>a</sup><br>Beta (95% CI) | *p* | Model 2<sup>b</sup><br>Beta (95% CI) | *p* | Model 3<sup>c</sup><br>Beta (95% CI) | *p* |
| --- | --- | --- | --- | --- | --- | --- |
| Impairment | 5.63 (4.03-7.23) | <.001 | 5.51 (3.91-7.11) | <.001 | 4.79 (3.21-6.37) | <.001 |
| Physical Activity | -0.03 (-0.19-0.13) | 0.72 | -0.04 (-0.20-0.12) | 0.62 | -0.01 (-0.17-0.15) | 0.90 |
| BMI | 1.03 (0.08-1.99) | 0.03 | 0.89 (-0.01-1.79) | 0.05 | 0.67 (-0.23-1.58) | 0.15 |
| FMI | 0.64 (-0.02-1.31) | 0.06 | 0.60 (-0.03-1.23) | 0.06 | 0.43 (-0.20-1.06) | 0.18 |
| Sleep Disturbances | 4.67 (2.14-7.20) | <.001 | 4.48 (1.96-7.01) | <.001 | 3.01 (0.58-5.45) | 0.02 |

*Note:* <sup>a</sup>Unadjusted model; <sup>b</sup>Adjusted for sex, age, SES, collection site, and single caregiver; <sup>c</sup>Adjusted for sex, age, SES, collection site, single caregiver and all diagnoses.

### 3.4 Supplementary Materials

Table S1 replicated the analyses reported in Table 2, though with odds ratios for PIU based on the parent-report version of the IAT instead of self-report. Self-report results for depression were replicated while those for ADHD combined type only approached significance. Of note, ASD emerged as a significant predictor of PIU. Tables S2 and S3 provide a complementary perspective of PIU, in which the presence of problematic behaviors is treated dimensionally. This was accomplished by utilizing multiple regression to examine the relationship between IAT total scores (self and parent report, respectively), as opposed to the dichotomous PIU variable, and psychopathology. Results for depression and ADHD combined type were replicated, and an additional significant relationship between ASD and PIU was observed. Social anxiety also emerged as a significant predictor, though less consistently so. The analyses reported in Table S4 replicated the associations with impairment reported in Table 3, though using parent-report versions of the IAT and CIS rather than self. Tables S5 and S6 were added to examine the relationship between PIU (measured using IAT total scores) and psychopathology using dimensional measures of symptomatology as opposed to DSM-5 diagnoses. For self-report, once all questionnaires were included as multiple predictors, only inattentive symptoms emerged as non-significant. For parent-report, once all questionnaires were included as multiple predictors for PIU, only inattention and depressive symptoms remained significant. Tables S7-S12 replicated all main analyses for both self and parent report with the maximum sample available for each analysis. Results obtained using the final sample (N = 564) were consistently replicated.

## 4. DISCUSSION

The current study examined associations between PIU and the presence of mental health disorders, impairment and health behaviors in a large, pediatric and transdiagnostic, self-referred community sample. We found consistent associations between PIU and depressive disorders and ADHD, combined type, based on both self- and informant-based reporting methods (see Tables S1, S3, S4, and S6), as well as whether psychopathology was measured using clinician diagnoses or dimensional report measures (see Tables S5 and S6). Importantly, PIU independently predicted impairment, even after accounting for the contributions of comorbid DSM-5 disorders. Consistent with prior studies,^36,50,53,73^ the presence of PIU was associated with increased sleep disturbances. Surprisingly, when compared with previous work,^69–71^ PIU was not consistently associated with physical fitness, though some results trended towards significance. This may be because some pediatric participants have more structured exercise and play times dictated by caregivers (e.g. Physical Education class, compulsory after-school sports, etc.) and thus the deleterious effects of PIU on physical fitness are prevented. Overall, findings from the present study suggest that PIU is associated with psychopathology, especially depression and ADHD, and may be associated with impairment even after accounting for comorbid psychopathology.

While depressive disorders have been consistently linked to PIU, associations with ADHD have been less commonly found.^39,45,51,53,84^ Caplan has suggested that depressed individuals tend to harbor negative views of their social competency, thus encouraging them to seek social encounters online, as these may be ‘easier’ to achieve since they allow for an increased flexibility in self-presentation through anonymity.^85^ Ko et al. have argued that internet use, particularly online gaming, provides necessary immediate reward, rapid response rates, and a plethora of activities to keep boredom at bay, thus making it highly appealing to individuals with ADHD.^32^ Additionally, poor self-control and high impulsivity also increase the risk of developing PIU in individuals with ADHD.^32^ Our findings have specific implications for the treatment of these disorders and the recognition of PIU, and may provide a pathway to understanding possible mechanisms for the maintenance of these disorders.

We did not confirm findings from earlier studies of an association between PIU with anxiety disorders and symptoms of anxiety.^47,50,55,57^ Researchers have theorized that individuals with anxiety, particularly social anxiety, use online relationships to compensate for poor real-life ones in a similar way as posited for individuals suffering from depression.^32,55^ However, many of these previous studies did not account for comorbidity with ADHD and depression.

In accordance with previous literature,^36,41,42,50,53,72,73^ we found an association between PIU and sleep problems, adding to growing awareness of the impact of inadequate sleep in U.S. adolescents on both mental and physical health.^86,87^ The persistence of this association after adjustment for not only demographic variables, but also ADHD and depression, suggests that internet use may directly influence sleep behavior or vice versa, thus providing some insight into the structure of the underlying relationships between PIU, psychopathology, and sleep.

Finally, it is worth noting that our findings imply that the effects of PIU on impairment are independent from the effects of comorbid psychopathology on impairment, contributing to the current debate on the structure of the relationship between impairment, PIU and psychopathology. The potential clinical implications suggest that PIU may not just be a worrisome behavior, but can indeed lead to serious impairment, especially for individuals who already suffer from a comorbid mental health disorder. With the recent addition of internet gaming disorder into the DSM-5, these considerations have begun to come into the light; future research should provide more information as to the specific negative consequences of different internet-related activities (i.e. sedentary behavior and sleep deprivation), and how these may contribute to impairment in various domains of life, particularly in populations at risk for psychiatric disorders.

There were several limitations to the current work. First, due to the community self-referred nature of the current sample, there is inherent selection bias that can limit generalizability due to the increased presence of psychopathology and other confounding factors. Second, PIU was only assessed through one questionnaire which, though demonstrated to be reliable, lacks specificity. The questionnaire does not specify what activities “internet use” and “being online” refer to, leaving interpretation to the respondent; it also has no mention of social media or smartphone usage, reflecting the timing of its creation. The Internet Process Addiction Test (IPAT)^88^ has been developed to address these issues, though is notably longer and yet to be heavily adopted in the literature. Third, while parent-reported PIU was available for children of all ages in the sample (5-21), self-reported questionnaires were only administered to children ages 7 and up. Fourth, the HBN sample is largely composed of individuals affected by one or more mental health or learning disorders, inherently increasing the likelihood of problematic behaviors among participants. As such, the odds ratios calculated for disorders such as ADHD and Depression may be an underestimate of what would be obtained in a more traditional, community representative sample. Finally, the sample size of the present work was not large enough to enable stable odds ratio estimates based on typically developing children alone.

Future research should focus on identifying the specific internet-related activities (i.e. social media, gaming, porn, gambling, etc^2^.) that are associated with different clinical disorders and comorbidity, as preliminary work has determined that different activities may have different relationships with psychopathology, behaviors, and personality characteristics^56,57,84,89–91^. Most importantly, potential mechanisms through which PIU is associated with impairment and sleep problems after adjustment for psychiatric disorders requires further inquiry. Identification of the directionality of these relationships should be pursued in prospective research in order to generate effective interventions to prevent the negative outcomes associated with PIU.

The present study provides empirical evidence for links between PIU and depressive disorders and Attention Deficit Hyperactivity Disorder in a large community sample of youth. The present study highlights the importance of exploring PIU in younger samples due to the myriad of negative behaviors associated with this construct. Further research is necessary to identify potential mechanisms for these associations and their impact on general functioning and sleep. It also presents options for interventions in order to weaken not only the occurrence of these negative behaviors but also the strength of the relationship between them.

## Data Availability

Data referred to in the manuscript was collected via the Healthy Brain Network research project.

https://www.nature.com/articles/sdata2017181

## ACKNOWLEDGEMENTS

We would like to thank the Healthy Brain Network team of research assistants and clinicians, as well as the families who participated in the study, for their hard work and dedication to making the project a reality, which not only enabled us to submit the present manuscript, but has provided a wealth of data that promises to enrich the field of pediatric psychiatry worldwide. The work presented here was primarily supported by gifts to the Child Mind Institute from Phyllis Green, Randolph Cowen, and Joseph Healey, as well as NIMH awards to Dr. Milham (U01MH099059, R01MH091864). This research was also supported by the Intramural Research Program of the NIMH (Merikangas) (grant number Z-01-MH002804). The views and opinions expressed in this article are those of the authors and should not be construed to represent the views of any of the sponsoring organizations, agencies, or U.S. Government.

